# The Effectiveness of Large Language Models in Providing Automated Feedback in Medical Imaging Education: A Protocol for a Systematic Review

**DOI:** 10.1101/2025.08.04.25332966

**Authors:** Mustafa Al-Mashhadani, Faika Ajaz, Shaista Salman Guraya, Farah Ennab

## Abstract

**Background:** Large Language Models (LLMs) represent an ever-emerging and rapidly evolving generative artificial intelligence (AI) modality with promising developments in the field of medical education. LLMs can provide automated feedback services to medical trainees (i.e. medical students, residents, fellows, etc.) and possibly serve a role in medical imaging education.

**Aim:** This systematic review aims to comprehensively explore the current applications and educational outcomes of LLMs in providing automated feedback on medical imaging reports.

**Methods:** This study employs a comprehensive systematic review strategy, involving an extensive search of the literature (Pubmed, Scopus, Embase, and Cochrane), data extraction, and synthesis of the data.

**Conclusion:** This systematic review will highlight the best practices of LLM use in automated feedback of medical imaging reports and guide further development of these models.

## 1. Introduction

### 1.1 Background

Medical imaging is a cornerstone of the modern medical environment, with reliable and accurate imaging attributed to improved patient outcomes (4). The availability of effective medical imaging has been shown to reduce mortality rates, improve diagnostic accuracy, and allow physicians a better understanding of patient presentations (3, 4). With these outcomes, medical imaging has become well integrated into the medical field. Thus, medical trainees (i.e., medical students, residents, fellows, continuing medical education) must be further educated in the context of imaging (1).

Additionally, artificial intelligence (AI) services in healthcare have demonstrated drastic improvements in capabilities and capacities in the medical field. AI has demonstrated an ability to improve diagnostic accuracy, reduce human error, lower healthcare costs, and enable personalised healthcare initiatives (3). The performance of Generative AI in medical imaging reports is continuously being studied. Generative AI can serve as an integrative part of the radiologists workflow, allowing for optimal reporting conditions, such as the prioritization of cases based on urgency and the reduction of limitations found in traditional reporting, through report standardization and the reduction of fatigue-induced errors (6).

For non-radiology-based applications, medical imaging education aids in anatomical knowledge retention while improving the identification of common pathologies and emergency cases (2). Non-radiology-based medical professionals and trainees must also be able to interpret and comprehend radiological reports, draw conclusions from these reports, and understand the principles of medical imaging studies, including how and when to request them (2). It is, therefore, paramount that medical education incorporates the element of medical imaging in educational curricula for all medical trainees.

In particular, the rise in popularity of Large Language Models (LLMs), like ChatGPT, within the context of medical education has revealed an emerging sector of automated and actionable feedback in imaging reports, providing real-time education for trainees (5). A preliminary search of the literature and review registration databases indicates a lack of literature reviews on the applications of LLMs in providing automated feedback to medical trainees in the context of medical imaging. Therefore, we are conducting a systematic review to explore and analyze the present literature and data regarding the effectiveness of LLMs in automated medical imaging feedback, identifying strengths and weaknesses of current models in this educational setting, while offering recommendations and initiatives for improving medical imaging education.

### 1.2 Study Aims

The aim of this systematic review is to explore the literature and identify how LLMs are being used to generate automated feedback on trainee reports during medical training.

### 1.3 Primary Objective

To examine the effectiveness of current LLM applications in providing feedback on medical imaging reports in medical educational settings.

### 1.4 Secondary Objectives

- Identify the quality of feedback LLMs generate (e.g., discrepancy detection, accuracy critique, language improvement, teaching points, etc.).
- Describe the educational levels and contexts in which LLMs are used (e.g., medical students, residents, fellows, continuing medical education, clinical rotations, simulation-based training, etc.).
- Explore the outcomes reported in studies evaluating LLM feedback use (e.g., diagnostic accuracy, accuracy compared to trained radiologists, trainee satisfaction, report quality, time saved, etc.).
- Evaluate the attitude and experiences of LLMs (e.g., expert validation, comparison to physician reports, user surveys, etc.).

## 2. Methodology

### 2.1 Systematic Review Framework

This methodology employs systematic search strategies. Articles were screened from four databases (PubMed, Scopus, Cochrane, and Embase) using Boolean operators based on a PICO framework, and incorporating relevant MeSH terms. A search strategy has been developed by two reviewers (as well as an expert health sciences librarian) alongside a pre-extraction protocol, both of which have been successfully registered on PROSPERO, a systematic review registration database.

In addition to the database search, a manual search and screening of references were conducted. Continuous manual searching will also occur throughout the study to ensure the inclusion of any newly published eligible studies. In the case of a scarcity of articles, the “hand-searching” technique will be utilized to identify additional relevant articles. A search of registration databases yielded no registrations for topics related to our study, and a search of grey literature alongside our continuous manual searching will allow us to draw a risk of bias assessment of our own research and whether we may have missed any studies.

The Covidence online platform was used for title and abstract screening, full-text review, data extraction, blinding the reviewers, and reporting results per the PRISMA 2020 guidelines checklist, ensuring comprehensive and transparent reporting of findings (7). Duplicates were removed automatically by Covidence, and a PRISMA flow chart was generated (7).

Data extraction was conducted by two blinded reviewers, with a final consensus occurring with a third reviewer present to resolve any conflicts. The study will also adhere to AMSTAR guidelines to ensure comprehensive, transparent, and reliable reporting of findings (8). Risk of bias for the included articles was assessed by two blinded reviewers utilizing the ROBINS-I bias assessment tool alongside reviewing funding sources and relevant conflicts of the included studies (9). The GRADE framework was also used to assess the certainty of our findings in the outcomes.

Moreover, the authors of this study have contacted the authors of any study for which we required clarification, and responses from other authors have been noted and taken into account for our research.

Any amendments to this protocol will be documented and updated on the PROSPERO registration page with justifications.

### 2.2 Study Selection Process

Articles that enter the screening phases (title-and-abstract and full-text reviews) must receive two “Yes” votes from two blinded reviewers, or one “Yes” and one “Maybe”, to move to full-text review. Articles receiving two “No” votes will be excluded, and any other combination will be placed into a “Conflict” section, where a third blinded reviewer will make the final decision.

### 2.3 Eligibility Criteria

#### 2.3.1 Table 1 - Inclusion Criteria

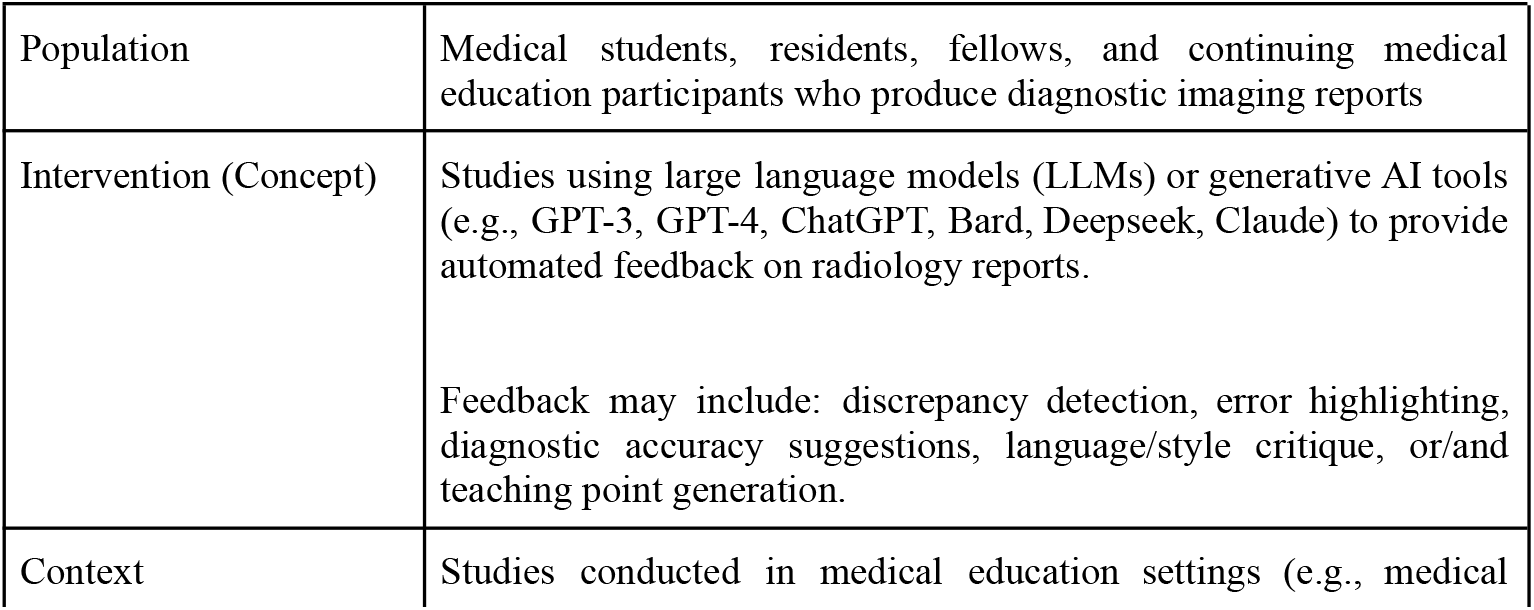
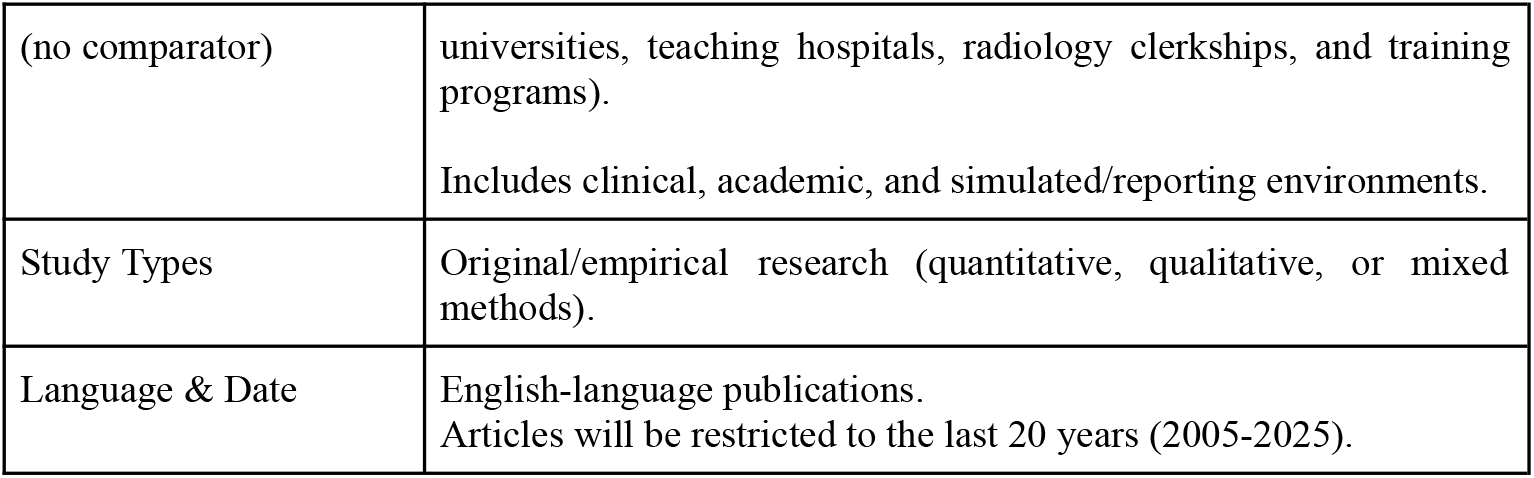

#### 2.3.2 Table 2 - Exclusion Criteria

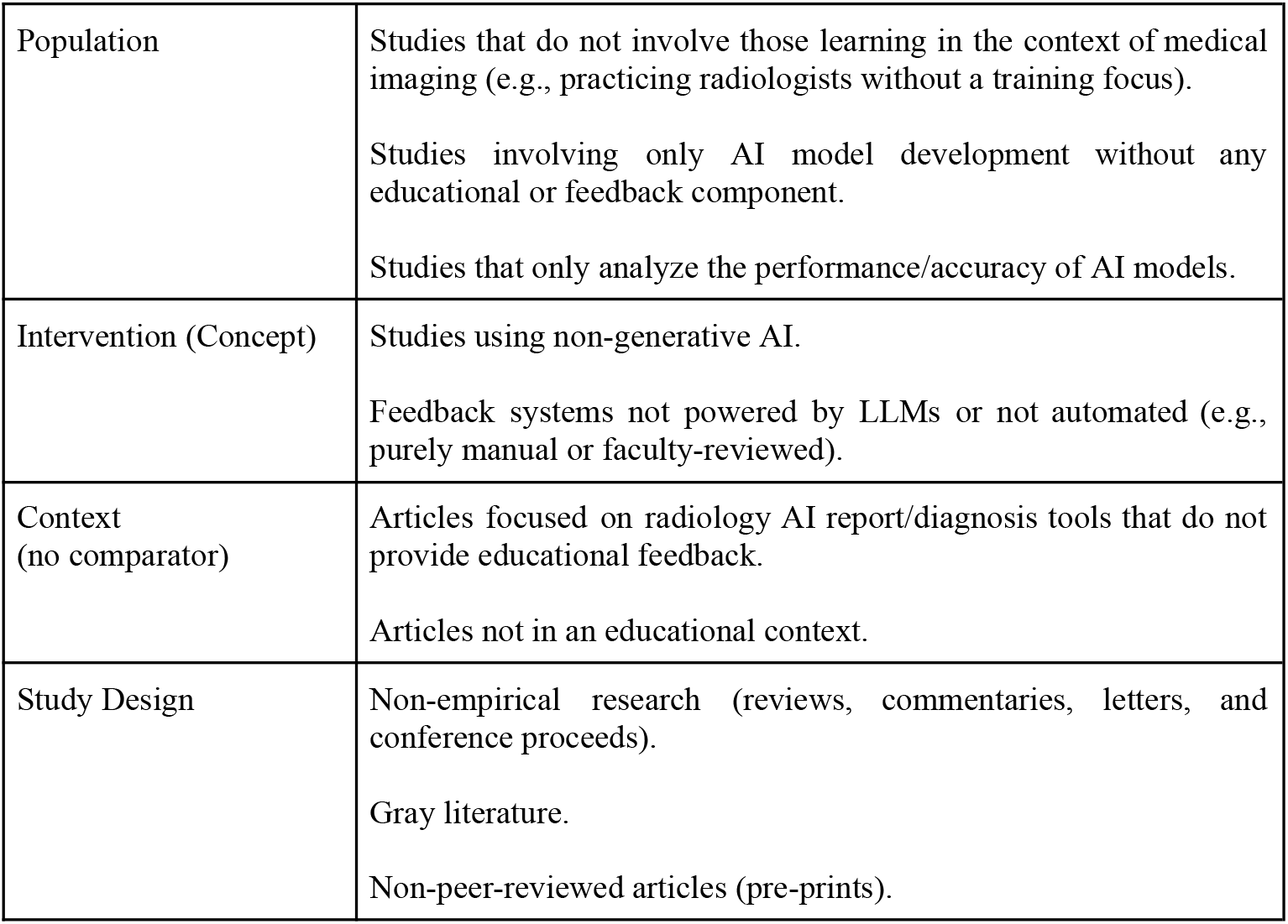

### 2.4 Search Strategy

#### 2.4.1 PICO Framework with MeSH Terminology

**Table 3:** PICO Framework Table

| PICO | Description | Keywords | MeSH Terms |
| --- | --- | --- | --- |
| Population | Radiology trainees in educational settings | radiology trainee, resident, medical student, fellow, CME, radiology education | Students, Medical; Education, Medical; Internship and Residency; Radiology; education |
| Intervention (Concept) | Use of large language models to provide automated feedback on radiology reports | large language models, LLMs, generative AI, ChatGPT, GPT-3, GPT-4, Claude, Bard, Gemini, automated feedback, AI feedback, DeepSeek | Natural Language Processing; Artificial Intelligence; Language Models |
| Context (no comparator) | Medical education environments (clinical or simulation) where trainees write diagnostic reports | hospital, radiology clerkship, simulation training, clinical rotation | Education, Medical, Undergraduate; Education, Medical, Graduate; Clinical Competence |
| Outcomes | Educational outcomes and quality of AI-generated feedback | diagnostic accuracy, discrepancy detection, satisfaction, report quality, training improvement | Feedback; Educational Measurement; Diagnostic Errors; Quality Improvement |

#### 2.4.2 Boolean Operators

**PubMed**

(

“Radiology”[MeSH] OR “Radiology Education” OR “Radiology Trainee” OR “Medical Student”[MeSH] OR resident OR fellow OR “Continuing Medical Education”)

AND

(“Artificial Intelligence”[MeSH] OR “Large Language Models” OR “Generative AI” OR “ChatGPT” OR “GPT-4” OR “Language Model” OR “Automated Feedback”)

AND

(“Education, Medical”[MeSH] OR “Education”[MeSH] OR “Clinical Training” OR “Simulation Training”)

AND

(“Feedback”[MeSH] OR “Educational Measurement”[MeSH] OR “Diagnostic Accuracy” OR “Discrepancy Detection” OR “Report Quality”

)

**Scopus (Operator 1)**

(

TITLE-ABS-KEY(“radiology trainee” OR “radiology trainees” OR “radiology resident” OR

“radiology residents” OR “medical student” OR “medical students” OR “clinical trainee” OR “clinical trainees” OR “subspecialty fellow” OR “fellow” OR “continuing medical education” OR “CME”)

)

AND

(

TITLE-ABS-KEY(“large language model” OR “large language models” OR “LLM” OR “LLMs” OR “generative AI” OR “artificial intelligence” OR “natural language processing” OR “language models” OR “ChatGPT” OR “GPT-3” OR “GPT-4” OR “Claude” OR “Bard” OR “Gemini” OR “Deepseek”)

)

AND

(

TITLE-ABS-KEY(“Radiology”)

)

AND

(

TITLE-ABS-KEY(“automated feedback” OR “feedback” OR “educational feedback” OR “discrepancy detection” OR “report quality” OR “language critique” OR “performance evaluation” OR “diagnostic accuracy” OR “teaching points” OR “Medical Education”)

)

**Scopus (Operator 2)**

(TITLE-ABS-KEY (radiology) AND TITLE-ABS-KEY (education) AND TITLE-ABS-KEY (artificial intelligence) AND TITLE-ABS-KEY (feedback))

**Embase**

(‘radiology/exp OR ‘radiology) AND ‘medical education:ti,ab,kw AND ‘artificial intelligence:ti,ab,kw

**Cochrane**

(“Radiology”):ti,ab,kw AND (“Education):ti,ab,kw AND (“Artificial Intelligence”):ti,ab,kw

- Note: PubMed & Scopus have a narrow search strategy yielding high results; however, Embase and Cochrane have a broad search strategy yielding low results. The authors were mindful of this, and thus, they used a refined search strategy specific to each database.

### 2.5 PRISMA Flow Chart

This systematic review extracted 831 studies from four databases (PubMed, Scopus, Embase, and Cochrane) on the 26^th^ of June, 2025. Covidence automatically removed 93 studies as they were duplicates. The authors conducted title-and-abstract screening on 738 studies, with 703 studies being excluded at this stage. All 35 articles that entered the full-text review were successfully retrieved for the full-text, and 7 articles were deemed to match the study inclusion criteria and contained no exclusion criteria. These 7 included articles were utilized in this systematic review to draw conclusions and further recommendations.

**Figure 1:**
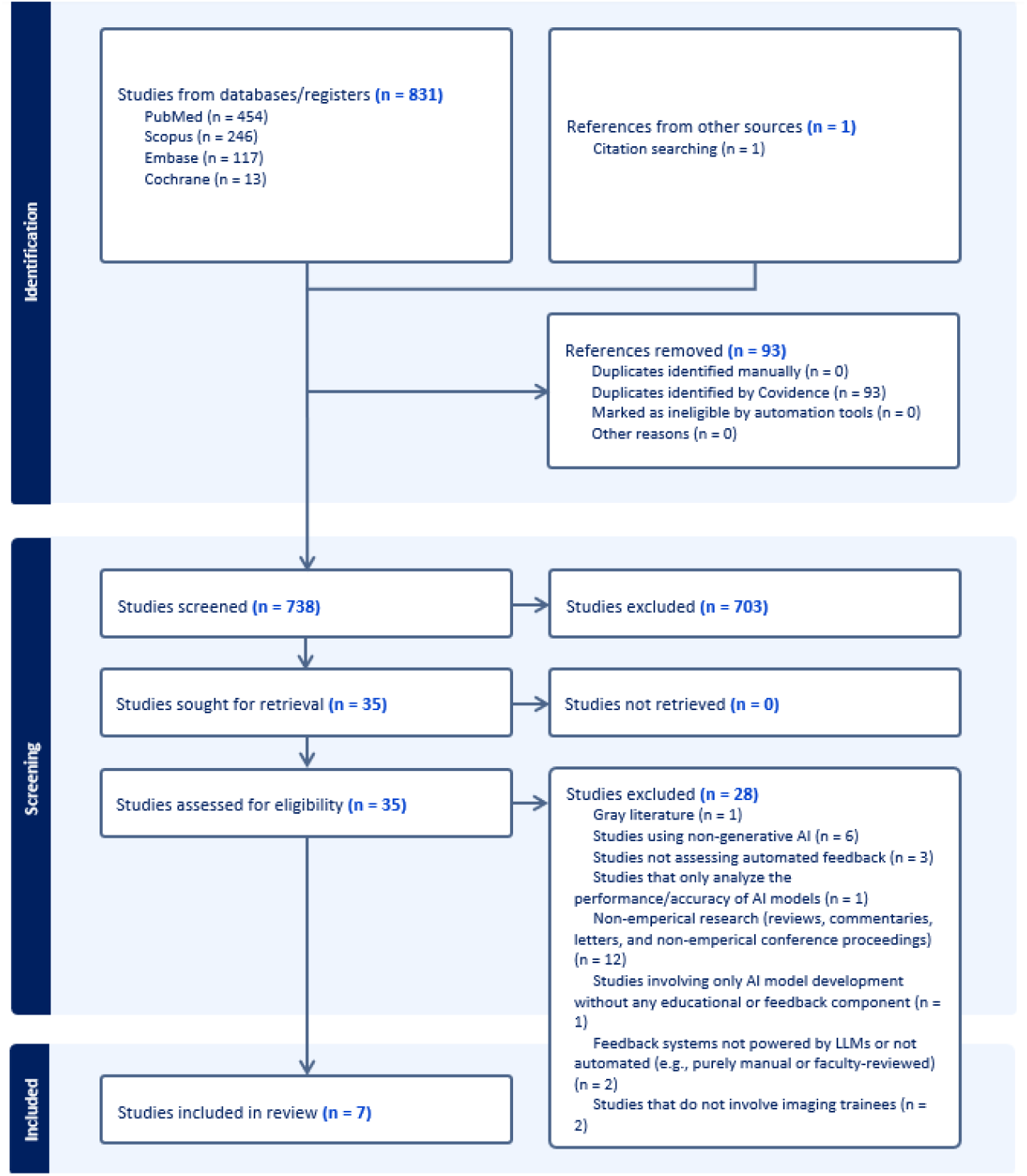
PRISMA flow diagram of this study’s screening and selection process.

## 3. Dissemination of Results

### 3.1 Ethics

Systematic reviews do not require formal ethical approval from an institutional board, as they do not involve direct active human engagement (primary data collection is not being performed). However, this systematic review aims to maintain the highest form of academic integrity, following guidelines and adhering to rigorous transparency and reliability protocols while ensuring correct handling of articles to prevent any plagiarism. Furthermore, we intend to be fully transparent in reporting our challenges and limitations while managing and declaring any conflicts of interest that may arise.

### 3.2 Dissemination Plans

The findings will be submitted to a high-impact and peer-reviewed journal for publication. Supplementary materials will also be shared and accessible to ensure wide dissemination of our findings and transparency in our work. A pre-extraction protocol has already been registered and is publicly available on PROSPERO, aiding in the early dissemination of our work. We also seek to publish a pre-print of our post-extraction protocol to raise awareness and gain engagement for our study before its final publication. Additionally, we aim to present the findings of this study in conferences as well to promote dissemination amongst researchers within relevant fields.

## 4. Data Extraction

### 4.1 Data Extraction

After data extraction, we have organised our findings into a series of summary tables. These tables will allow us to assess the effectiveness of feedback generated by large language models in medical imaging education, especially in the feedback process on trainee reports.

We aim to identify:

- Which models are used most frequently and in what context?
- The types of feedback they provide.
- The contexts in which they’re deployed (i.e. simulation vs. clinical practice).
- The types of trainees involved (medical students, residency, fellowship, CME, etc.).
- The outcomes of the feedback.

### 4.2 Data Extraction Table Example

**Table 4.** Data Extraction Table Design Example.

| Category | Details to Extract |
| --- | --- |
| Study | Author(s), year of publication. |
| Country | Country where the study was conducted. |
| Trainee Level | Medical students, residents, fellows, CME. |
| Clinical Setting | Teaching hospital, simulation center, academic institution. |
| Large Language Model (LLM) | Specific models used (e.g., ChatGPT, GPT-4, Bard, Gemini, DeepSeek, Claude, etc.). |
| Feedback Function | Type of feedback (e.g. discrepancy detection, teaching points, language critique, etc.). |
| Educational Context | Clinical rotations, clerkships, simulated reporting, structured training programs. |
| Study Design | Observational, experimental, qualitative, quantitative, mixed-methods. |
| Sample Size | Number of participants or trainee reports analyzed. |
| Key Outcomes | Diagnostic accuracy, Time saved, Satisfaction, Report quality, Comparison to expert. |
| Conclusions / Recommendations | Summary of key insights, lessons learned, future recommendations. |

### 4.3 Data Synthesis, Results, and Discussion

Extracted data has been organised in tables summarizing study characteristics (e.g., author, year, country), trainee level, clinical setting, type of large language model used, type of feedback, educational context, study design, sample design, key outcomes, and conclusions/recommendations.

The results section will present a structured overview of how LLMs are being used to provide automated feedback on medical imaging reports, highlighting trends, tools, and outcomes. We will present our narrative synthesis, categorised by intervention (LLM) and educational outcomes. Study characteristics and key findings will be summarized in tables. We will report common themes, contextual variations, and literature gaps across the included studies.

In the discussion section, we will interpret these findings in relation to current radiological educational practices. We will examine the implications for training programs and offer recommendations for future research and development. We will also discuss limitations and methodological rigour of the included studies and of the review process itself.

## Conclusion

This systematic review will provide a comprehensive overview of the literature, exploring applications of generative AI, specifically LLMs, in medical imaging educational settings. This study will provide further recommendations to enhance medical education through the use of continually advancing generative AI models. It will also guide medical educators in the implementation of LLMs in developing tools and mechanisms for medical institutions, while also identifying areas for future research into the applications of generative AI.

## Data Availability

All data used in this study were derived from previously published sources. No new public datasets were generated or analyzed. Access to the original sources can be obtained through their respective publications.

## Appendix

### Prospective Registration

This study was successfully registered prospectively on PROSPERO on the 26^th^ of June, 2025. The PROSPERO registration code is: CRD420251081394, available from https://www.crd.york.ac.uk/PROSPERO/view/CRD420251081394

## Availability of Code, Tables, and Data

Our data set and the tools we used to extract it will be available as supplementary material in our final publication.

